# Development and Validation of a Nomogram for Predicting Survival in Patients With Cardiogenic Shock

**DOI:** 10.1101/2024.11.14.24317361

**Authors:** Dingfeng Fang, Huihe Chen, Hui Geng, Xiahuan Chen, Meilin Liu

## Abstract

**Background:** There is currently a lack of easy-to-use tools for assessing the severity of cardiogenic shock (CS) patients. This study aims to develop a nomogram for evaluating severity in CS patients regardless of the underlying cause.

**Methods and Results:** The MIMIC-IV database was used to identify 1923 CS patients admitted to the ICU. A multivariate Cox model was developed in the training cohort (70%) based on LASSO regression results. Factors such as age, systolic blood pressure, arterial oxygen saturation, hemoglobin, serum creatinine, blood glucose, arterial pH, arterial lactate, and norepinephrine use were incorporated into the final model. This model was visualized as a Cardiogenic Shock Survival Nomogram (CSSN) to predict 30-day survival rates. The model’s c-statistic was 0.75 (95% CI 0.73–0.77) in the training cohort and 0.73 (95% CI 0.70– 0.77) in the validation cohort, demonstrating good predictive accuracy. The AUC of the CSSN for 30-day survival probabilities was 0.76 in the training cohort and 0.73 in the validation cohort. Calibration plots showed strong concordance between predicted and actual survival rates, and decision curve analysis (DCA) affirmed the model’s clinical utility. The CSSN outperformed the Cardiogenic Shock Score (CSS) in various metrics, including c-statistic, time-dependent ROC, calibration plots, and DCA (c-statistic: 0.75 versus 0.72; AUC: 0.76 versus 0.73). Sensitivity analyses confirmed the model’s robustness across both AMI-CS and non-AMI-CS subgroups.

**Conclusions:** The CSSN was developed to predict 30-day survival rates in CS patients irrespective of the underlying cause, showing good performance and potential clinical utility in managing CS.

**CLINICAL PERSPECTIVE:** **What Is New?**

1. This study presents the development and validation of a novel nomogram, the Cardiogenic Shock Survival Nomogram (CSSN), specifically designed for predicting 30-day survival rates in patients with cardiogenic shock (CS), regardless of the underlying cause.
2. Utilizing a large cohort from the MIMIC-IV database, this research integrates multiple available clinical factors—such as age, systolic blood pressure, arterial oxygen saturation, hemoglobin levels, serum creatinine, blood glucose, arterial pH, arterial lactate, and norepinephrine use—into a convenient and easy to use the prognostic tool.

**What Are the Clinical Implications?**

1. Enhanced Decision-Making: Clinicians can utilize the CSSN as a practical tool to assess the severity and prognosis of CS patients, enabling more informed decision-making regarding treatment strategies.
2. Improved Risk Stratification: By incorporating easily obtainable clinical parameters such as age, systolic blood pressure, arterial oxygen saturation, and biochemical markers, the CSSN provides a straightforward method for risk stratification, facilitating the identification of high-risk patients who may benefit from more intensive monitoring or advanced therapeutic interventions.

## Introduction

Cardiogenic shock (CS) is a life-threatening medical emergency with a high mortality rate of 40% to 60%, despite advancements in medical care, leading to prolonged suffering for affected patients and significant healthcare costs^1–4^. The causes and severity of CS can vary widely, which results in different treatment approaches and prognoses^3,5,6^. Early identification of high-risk patients can help promptly implement reasonable and practical treatment measures. Thus, assessing the severity of the condition is essential, as it directly influences clinical decisions, such as whether to administer mechanical circulatory support (MCS). Several risk assessment tools for CS have been developed, with the most widely accepted being the IABP-SHOCK II Score^7^, CardShock risk Score^8^, and Cardiogenic Shock Score (CSS)^9^. However, these models are primarily presented as scoring systems, which are relatively complex and thus less frequently used in clinical practice. Nomograms are graphical representations of mathematical models employed to forecast outcomes by assessing clinical events and incorporating key prognostic factors across various diseases^10,11^. They offer intuitive, fast, straightforward, and user-friendly advantages. Currently, no predictive model for CS uses a nomogram presentation available for clinical use.

Moreover, the SHOCK II Score and CardShock Risk Score mainly target the AMI-CS population, which may not apply to non-AMI-CS patients. Recent epidemiological studies have found that AMI as a cause of CS has decreased to approximately 30%, indicating that AMI is no longer the predominant cause of cardiogenic shock^3,12^. The Society for Cardiovascular Angiography and Intervention (SCAI) classification appears to be the most widely used risk assessment tool, but it is more focused on staging the progression of CS rather than providing a specific and easy-to-use risk scoring system^6,13^. Given these reasons, our study aims to develop an easy-to-use predictive model for CS, irrespective of the underlying cause, in the emergency setting, which accounts for survival assessments. This model will be presented as a Cardiogenic Shock Survival Nomogram (CSSN) to enhance clinical applicability. The establishment of this model will aid physicians in timely assessing the severity of CS, thereby making appropriate clinical decisions to improve patient survival rates.

## Methods

### Data Source and Study Participants

This study utilized the MIMIC-IV database, a comprehensive repository of critical care data^14–16^. MIMIC-IV is a collection of clinical records from patients treated in intensive care units (ICU) at the Beth Israel Deaconess Medical Center. The first author of this study, Dingfeng Fang, has completed the Collaborative Institutional Training Initiative (CITI) program course of Massachusetts Institute of Technology Affiliates (Human Research, Data or Specimens Only Research, and Refresher Course), granting access to the MIMIC-IV database (Record ID: 50924352). Data retrieval from the database was conducted using Structured Query Language (SQL). For this study, all patients who experienced cardiogenic shock (CS) during their hospital stay and subsequently received further treatment in the ICU were included. Patients for this cohort were identified by querying the MIMIC-IV database for instances of the International Classification of Diseases, Ninth Revision (ICD-9) code 785.51, and the Tenth Revision (ICD-10) code R57.0, both denoting cardiogenic shock. To reduce survival and treatment biases caused by readmissions, only patients admitted to the ICU for the first time were included in the analysis.

### Data Collection

Demographic, vital signs, laboratory, clinical, and outcome data, procedures, and therapies performed during the ICU and hospital stay were collected from the MIMIC-IV database. Radiographic, invasive hemodynamic, and physical examination data were not available. Baseline characteristics included gender, age, admission diagnosis (CS, AMI), comorbidities (previous MI, hypertension, diabetes, renal impairment), and smoking history. Systolic blood pressure (SBP), diastolic blood pressure (DBP), mean arterial pressure (MAP), arterial oxygen saturation (SpO_2_), and partial pressure of oxygen (PaO_2_) were all recorded from the lowest values within the first 24 hours of admission. The serum creatinine level was recorded as the maximum value within the first 24 hours of admission. Other laboratory tests were defined by the first recorded value after ICU admission or the value closest to ICU admission. Laboratory tests included blood assays (hemoglobin, leukocytes, platelets), arterial blood gas analysis (pH, PaO_2_, partial pressure of carbon dioxide (PaCO_2_), lactate, base excess, total carbon dioxide), coagulation profile, liver function, renal function, serum albumin, electrolyte measurements, blood glucose, low-density lipoprotein cholesterol (LDL-C), and cardiac enzymes (troponin T, N-terminal pro-b-type natriuretic peptide (NT-proBNP)). Critical respiratory and cardiovascular treatments administered to the patients, such as norepinephrine, dopamine, dobutamine, epinephrine infusions, invasive mechanical ventilation, IABP, and continuous renal replacement therapy (CRRT), were also recorded. The primary outcome was all-cause mortality within 30 days. The database determined the date of death based on state records and hospital documentation. In cases where data from both sources are available, the hospital records take precedence.^14–16^ All patients had access to complete follow-up data for one year. Survival duration was calculated from the time of hospital admission until death.

### Statistical Analysis

Data analysis was performed using R version 4.3.3. A significance level of less than 0.05 was considered statistically significant for all analyses. Variables with more than 20% missing values were excluded. For variables with less than 20% missing values, missing data were imputed using multiple imputations with random forests, implemented through the mice package in R. The data were randomly split into training (70%) and validation (30%) sets for subsequent analysis. Continuous variables were reported as the median and interquartile range (IQR), while categorical variables were presented as frequencies and percentages. Differences between groups were compared using chi-square tests for categorical variables and Wilcoxon rank-sum tests for continuous variables.

In the training cohort, LASSO regression was employed to identify predictors of 30-day mortality in patients with CS. Potential variables for model construction were based on readily available point-of-care parameters and previous studies. The most parsimonious set of variables selected by LASSO regression was used to develop a multivariate Cox proportional hazards model. Multicollinearity was evaluated using the variance inflation factor (VIF). Harrell’s Concordance Index (c-statistic) assessed the model’s predictive accuracy. Factors with prognostic significance in the multivariate Cox regression analysis were used to construct 30-day survival prediction models (1 - mortality), visualized using a nomogram. The nomogram predicted the 30-day mortality risk for all patients, categorizing them into four risk groups: low risk (0%-15%), moderate risk (16%-49%), high risk (50%-84%), and very high risk (85%-100%). Kaplan-Meier plotter was used to perform 1-year survival analysis for the four patient groups.

Time-dependent Receiver Operating Characteristic (ROC) curves and Area Under the Curve (AUC) were used to evaluate the discriminative power of the nomogram. Calibration curves assessed the difference between the actual and nomogram-predicted event-free survival rates using bootstrapping (1000 resamplings). The multivariate Cox proportional hazards model was similarly established in the validation cohort, and the c-statistic was calculated to evaluate its predictive accuracy. In the validation cohort, the performance of the nomogram was similarly evaluated using ROC curves, AUC, and calibration curves. Finally, decision curve analysis (DCA) was conducted to evaluate the clinical utility of the prediction models over 30 days. DCA assesses the net benefit across a range of threshold probabilities, comparing the benefits of correctly identifying high-risk patients with the harms of unnecessary interventions in low-risk patients, helping to determine the optimal decision threshold. Additionally, this study compared the accuracy and discrimination of our model with that of the Cardiogenic Shock Score (CSS).

Sensitivity analyses were conducted in two ways: (1) performing multivariate Cox regression analysis using the variables from the nomogram on all AMI-CS patients and calculating the c-statistic and AUC to evaluate the predictive accuracy in AMI-CS patients; (2) performing multivariate Cox regression analysis using the variables from the nomogram on all non-AMI-CS patients and calculating the c-statistic and AUC to evaluate its predictive accuracy in non-AMI-CS patients.

## Results

### Study Population and Baseline Characteristics

A total of 2216 patients with admission diagnosis of CS based on ICD-9 code 785.51 and ICD-10 code R57.0. Of these, 152 were readmissions, and 141 did not require ICU admission; these patients were excluded from this study. The final cohort included 1923 patients, with 1346 randomly assigned to the training cohort and 577 to the validation cohort. Variables with over 20% missing data (albumin, troponin T, NT-proBNP, D-dimer, and LDL-C) were excluded. Variables with less than 20% missing data were imputed using the ‘mice’ package in R, utilizing random forest-based multiple imputation, resulting in 89 imputed datasets.

Of the 1923 patients, the median age was 70 (IQR 60–79), and 40.46% were female. Acute myocardial infarction-related cardiogenic shock (AMI-CS) was present in 28.13% of patients. Medical history included 14.25% of patients with previous myocardial infarction, 26.94% with hypertension, 40.04% with diabetes, and 46.33% with renal impairment. During the first 24 hours after ICU admission, the median minimum values for key vital signs were 79 mmHg (IQR 68.75-87) for SBP and 91% (IQR 87-94%) for SpO2. Abnormal laboratory findings included a median, minimum oxygen partial pressure of 46 mmHg (IQR 33-79), a median maximum serum creatinine of 159.12 µmol/L (IQR 106.08-247.52), and a median arterial lactate level of 3.3 mmol/L (IQR 1.90-6.40 mmol/L). Arterial blood gas pH was below 7.0 in 81 patients (4.21%).

### Treatment and Outcome

As shown in Table 1, patients received various treatments during hospitalization, including vasopressors, respiratory and circulatory support, and renal replacement therapy. Treatments included dopamine (n=465, 24.18%), dobutamine (n=481, 25.01%), norepinephrine (n=1351, 70.25%), epinephrine (n=469, 24.39%), intra-aortic balloon pump (IABP) (n=319, 16.59%), mechanical ventilation (n=1111, 57.77%), and continuous renal replacement therapy (CRRT) (n=306, 15.91%). During the one-month follow-up, 1097 patients survived (57.05%), and 826 patients died (42.95%).

**Table 1.** Baseline characteristics, treatment, and outcomes of the study population.

| **Group** | **All** | **Training cohort** | **Validation cohort** | **P\*** |
| --- | --- | --- | --- | --- |
| Number | 1923 | 1346 | 577 |  |
| Age, year | 70.00 (60.00-79.00) | 70.00 (60.00-78.75) | 71.00 (60.00-80.00) | 0.179 |
| Female | 778 (40.46%) | 540 (40.12%) | 238 (41.25%) | 0.644 |
| **Etiology** |  |  |  |  |
| AMI |  |  |  | 0.712 |
| non-AMI | 1382 (71.87%) | 960 (71.32%) | 422 (73.14%) |  |
| NSTEMI | 406 (21.11%) | 289 (21.47%) | 117 (20.28%) |  |
| STEMI | 135 (7.02%) | 97 (7.21%) | 38 (6.59%) |  |
| Cardiac arrest, n(%) | 238 (12.38%) | 163 (12.11%) | 75 (13.00%) | 0.588 |
| **Comorbidities** |  |  |  |  |
| Previous MI, n(%) | 274 (14.25%) | 184 (13.67%) | 90 (15.60%) | 0.268 |
| Hypertension, n(%) | 518 (26.94%) | 363 (26.97%) | 155 (26.86%) | 0.962 |
| Diabetes, n(%) | 770 (40.04%) | 540 (40.12%) | 230 (39.86%) | 0.916 |
| Renal impairment, n (%) | 891 (46.33%) | 619 (45.99%) | 272 (47.14%) | 0.642 |
| Smoker, n (%) | 206 (10.71%) | 141 (10.48%) | 65 (11.27%) | 0.608 |
| **Vital signs** |  |  |  |  |
| SBP, mmHg | 79.00 (68.75-87.00) | 79.00 (68.12-87.00) | 79.00 (69.00-87.00) | 0.57 |
| DBP, mmHg | 39.00 (31.00-46.00) | 39.75 (31.00-46.00) | 39.00 (31.00-46.00) | 0.675 |
| MAP, mmHg | 51.00 (42.00-59.00) | 51.00 (42.00-59.00) | 51.00 (42.00-58.50) | 0.418 |
| SpO2, % | 91.00 (87.00-94.00) | 92.00 (87.25-94.00) | 91.00 (86.00-94.00) | 0.105 |
| GCS | 15.00 (15.00-15.00) | 15.00 (15.00-15.00) | 15.00 (15.00-15.00) | 0.759 |
| **Laboratory findings at admission** |  |  |  |  |
| Hemoglobin, g/dL | 11.20 (9.50-12.85) | 11.20 (9.60-12.90) | 11.10 (9.30-12.80) | 0.226 |
| Aspartate aminotransferase, U/L | 70.00 (32.00-245.00) | 70.50 (32.00-238.00) | 69.00 (32.00-270.00) | 0.993 |
| Alanine aminotransferase, U/L | 45.00 (22.00-149.50) | 45.00 (21.25-147.50) | 44.00 (22.00-151.00) | 0.869 |
| Serum creatine, umol/L | 159.12<br>(106.08-247.52) | 159.12<br>(106.08-238.68) | 159.12<br>(106.08-247.52) | 0.804 |
| Serum sodium, mmol/L | 138.00<br>(134.00-141.00) | 138.00<br>(134.00-141.00) | 137.00<br>(135.00-140.00) | 0.662 |
| Serum potassium, mmol/L | 4.30 (3.80-4.80) | 4.30 (3.80-4.80) | 4.20 (3.80-4.80) | 0.302 |
| Serum calcium, mg/dL | 8.50 (7.90-8.90) | 8.50 (7.90-8.90) | 8.50 (8.00-8.90) | 0.925 |
| Blood glucose |  |  |  | 0.281 |
| <11.1 mmol/L | 1452 (75.51%) | 1007 (74.81%) | 445 (77.12%) |  |
| ≥11.1 mmol/L | 471 (24.49%) | 339 (25.19%) | 132 (22.88%) |  |
| **Arterial blood gas** |  |  |  |  |
| pH |  |  |  | 0.228 |
| <7.0 | 81 (4.21%) | 55 (4.09%) | 26 (4.51%) |  |
| 7.0-7.35 | 1310 (68.12%) | 933 (69.32%) | 377 (65.34%) |  |
| >7.35 | 532 (27.67%) | 358 (26.60%) | 174 (30.16%) |  |
| PaO<sub>2</sub>, mmHg | 46.00 (33.00-79.00) | 46.00 (33.00-79.00) | 47.00 (33.00-79.00) | 0.833 |
| PaCO<sub>2</sub>, mmHg | 40.00 (34.00-47.00) | 41.00 (35.00-47.75) | 39.00 (34.00-46.00) | 0.049 |
| lactate, mmol/L | 3.30 (1.90-6.40) | 3.40 (2.00-6.50) | 3.00 (1.90-5.80) | 0.068 |
| **Treatments** |  |  |  |  |
| Dopamine, n (%) | 465 (24.18%) | 318 (23.63%) | 147 (25.48%) | 0.385 |
| Dobutamine, n (%) | 481 (25.01%) | 356 (26.45%) | 125 (21.66%) | 0.026 |
| Norepinephrine, n (%) | 1351 (70.25%) | 960 (71.32%) | 391 (67.76%) | 0.118 |
| Epinephrine, n (%) | 469 (24.39%) | 339 (25.19%) | 130 (22.53%) | 0.214 |
| IABP, n (%) | 319 (16.59%) | 227 (16.86%) | 92 (15.94%) | 0.619 |
| Invasive ventilation, n (%) | 1111 (57.77%) | 792 (58.84%) | 319 (55.29%) | 0.148 |
| CRRT, n (%) | 306 (15.91%) | 219 (16.27%) | 87 (15.08%) | 0.512 |
| **Outcomes** |  |  |  |  |
| Death during 1-month | 826 (42.95%) | 591 (43.91%) | 235 (40.73%) | 0.197 |
| Death during 1-year | 1106 (57.51%) | 776 (57.65%) | 330 (57.19%) | 0.852 |

Data are presented as median (interquartile range) unless otherwise indicated.
Abbreviation: AMI, acute myocardial infarction; Previous MI, previous myocardial infarction; NSTEMI, non-ST-elevation myocardial infarction; STEMI, ST-Elevation Myocardial Infarction. SBP, systolic blood pressure; DBP, diastolic blood pressure; MAP, mean arterial pressure; SPO<sub>2</sub>, arterial oxygen saturation; GCS, Glasgow Coma Scale; PaO<sub>2</sub>, Partial pressure of oxygen; PaCO<sub>2</sub>, partial pressure of carbon dioxide; IABP, intra-aortic balloon pump; CRRT, continuous renal replacement therapy.
\* Comparison between the training and validation cohort.

### Predictor Selection and Nomogram Construction

LASSO regression was used to identify predictors of 30-day mortality in CS patients from the training cohort (Figure 1). Potential variables for model construction were based on readily available point-of-care parameters and previous studies, including gender, age, admission diagnosis of AMI, admission of cardiac arrest, comorbidities (previous MI, hypertension, diabetes, renal impairment), smoking history, SBP, heart rate, SpO2, Glasgow Coma Scale (GCS), hemoglobin, alanine aminotransferase (ALT), aspartate aminotransferase (AST), serum creatinine, serum sodium, serum potassium, serum calcium, serum chloride, blood glucose, arterial blood gas (pH, partial pressure of oxygen, lactate), and treatments (dopamine, dobutamine, norepinephrine, epinephrine, IABP, invasive ventilation, CRRT). The lambda.1se (λ = 0.0621) was used to obtain the simplest model with nine predictors: Age, SBP, SpO_2_, hemoglobin, serum creatinine, blood glucose, pH, arterial lactate, and norepinephrine. These predictors were used to develop a multivariate Cox proportional hazards model, and the result is shown in Table 2. Higher age, serum creatinine, blood glucose, arterial lactate, norepinephrine use, and lower SBP, SpO2, hemoglobin, and pH were associated with increased mortality. Multicollinearity was assessed, and variance inflation factor (VIF) values were below the threshold, indicating no significant multicollinearity issues. The model demonstrated good discriminative ability, as evidenced by a c-statistic of 0.75 (95% CI 0.73– 0.77) and an area under the curve (AUC) of 0.76. Additionally, external validation using CSS data from the training set yielded a c-statistic of 0.72 (95% CI 0.70–0.74) with an AUC of 0.73. A nomogram was constructed using the multivariate Cox model to predict 30 survival rates (1 - mortality rate) (Figure 2).

**Figure 1.**
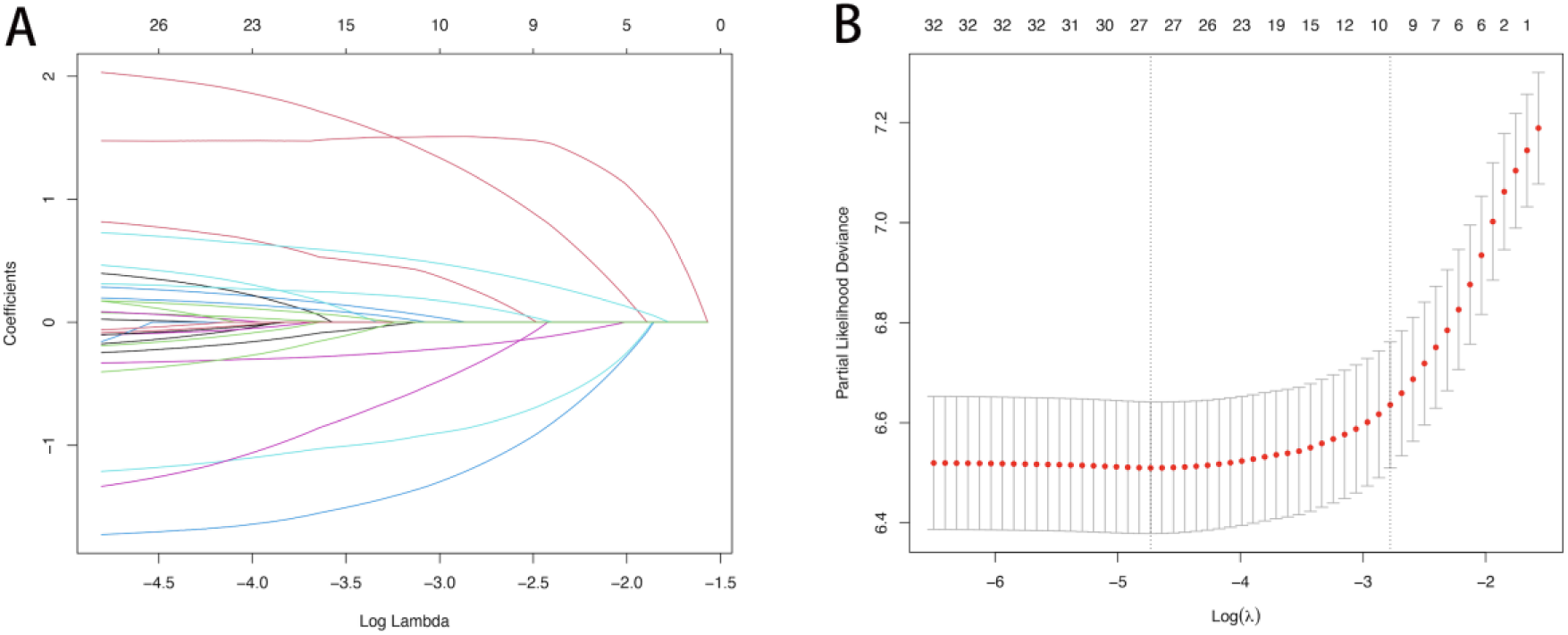
LASSO regression analysis of predictive factors for 30-day mortality of patients with CS.

**Figure 2.**
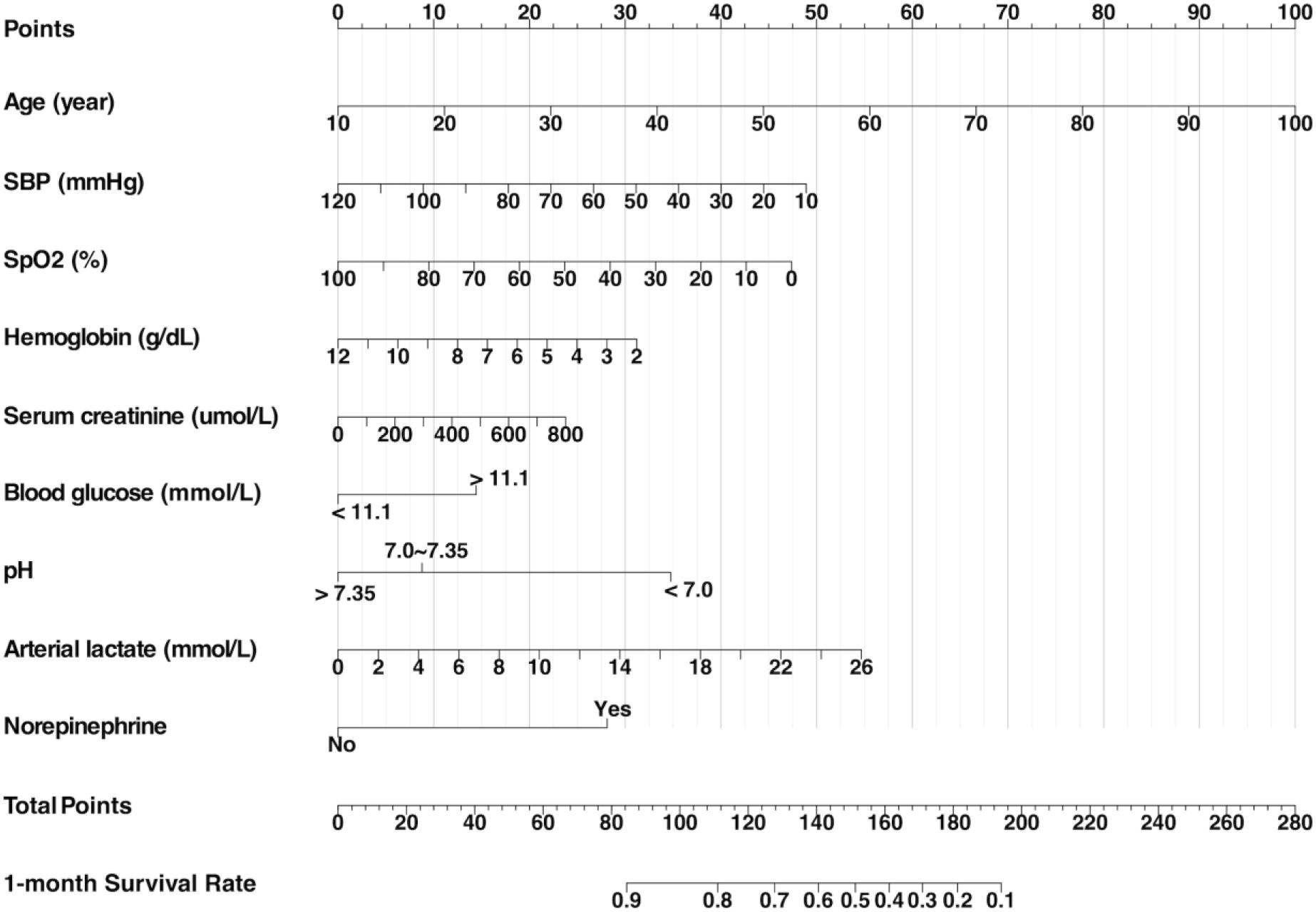
Cardiogenic Shock Survival Nomogram (CSSN). The nomogram predicts the 30-day survival rates for patients with CS. Abbreviations: SBP, systolic blood pressure; SpO_2_, peripheral oxygen saturation.

**Table 2.** Multivariate Cox analysis of potential prognostic factors identified by LASSO regression.

| **Variables** | **VIF** | **Coefficients** | **HR (95%CI)** | **P** |
| --- | --- | --- | --- | --- |
| **Age, year** | 1.042 | 0.031 | 1.032 (1.025, 1.039) | <0.0001 |
| **SBP, mmHg** | 1.285 | -0.013 | 0.988 (0.982, 0.993) | <0.0001 |
| **SpO<sub>2</sub>, %** | 1.263 | -0.013 | 0.987 (0.981, 0.993) | <0.0001 |
| **Hemoglobin, g/dL** | 1.019 | -0.088 | 0.916 (0.871, 0.964) | 0.0007 |
| **Serum creatinine, umol/L** | 1.096 | 0.001 | 1.001 (1.001, 1.002) | 0.0024 |
| **Blood glucose** | 1.098 |  |  |  |
| **<11.1 mmol/L** |  | Reference |  |  |
| **≥11.1mmol/L** |  | 0.406 | 1.501 (1.254, 1.796) | <0.0001 |
| **pH (Arterial blood gas)** | 1.455 |  |  |  |
| **<7.1** |  | Reference |  |  |
| **7.1~7.35** |  | -0.731 | 0.481 (0.342, 0.677) | <0.0001 |
| **>7.35** |  | -0.978 | 0.376 (0.249, 0.567) | <0.0001 |
| **Arterial lactate, mmol/L** | 1.657 | 0.059 | 1.061 (1.038, 1.084) | <0.0001 |
| **Norepinephrine** | 1.062 | 0.791 | 2.205 (1.747, 2.74) | <0.0001 |

This multivariate Cox proportional hazards model was performed to predict 30-day mortality in patients with CS in the training cohort (N=1346). The Harrell's concordance index (c-statistic) is 0.75 (95% CI 0.73–0.77).
Multicollinearity among variables was evaluated by calculating the variance inflation factor (VIF). A VIF greater than 10 indicates the presence of multicollinearity.
Abbreviations: HR, hazard ratio; CI, confidence interval; SBP, systolic blood pressure; SpO<sub>2</sub>, peripheral oxygen saturation.

The nomogram stratified all patients into four risk categories for 30-day mortality: low risk (0%–15%), moderate risk (16%–49%), high risk (50%–84%), and very high risk (85%– 100%). Using Kaplan-Meier curves, a one-year survival analysis was conducted across these four groups, revealing significant differences in mortality within the first year (Figure 3).

**Figure 3.**
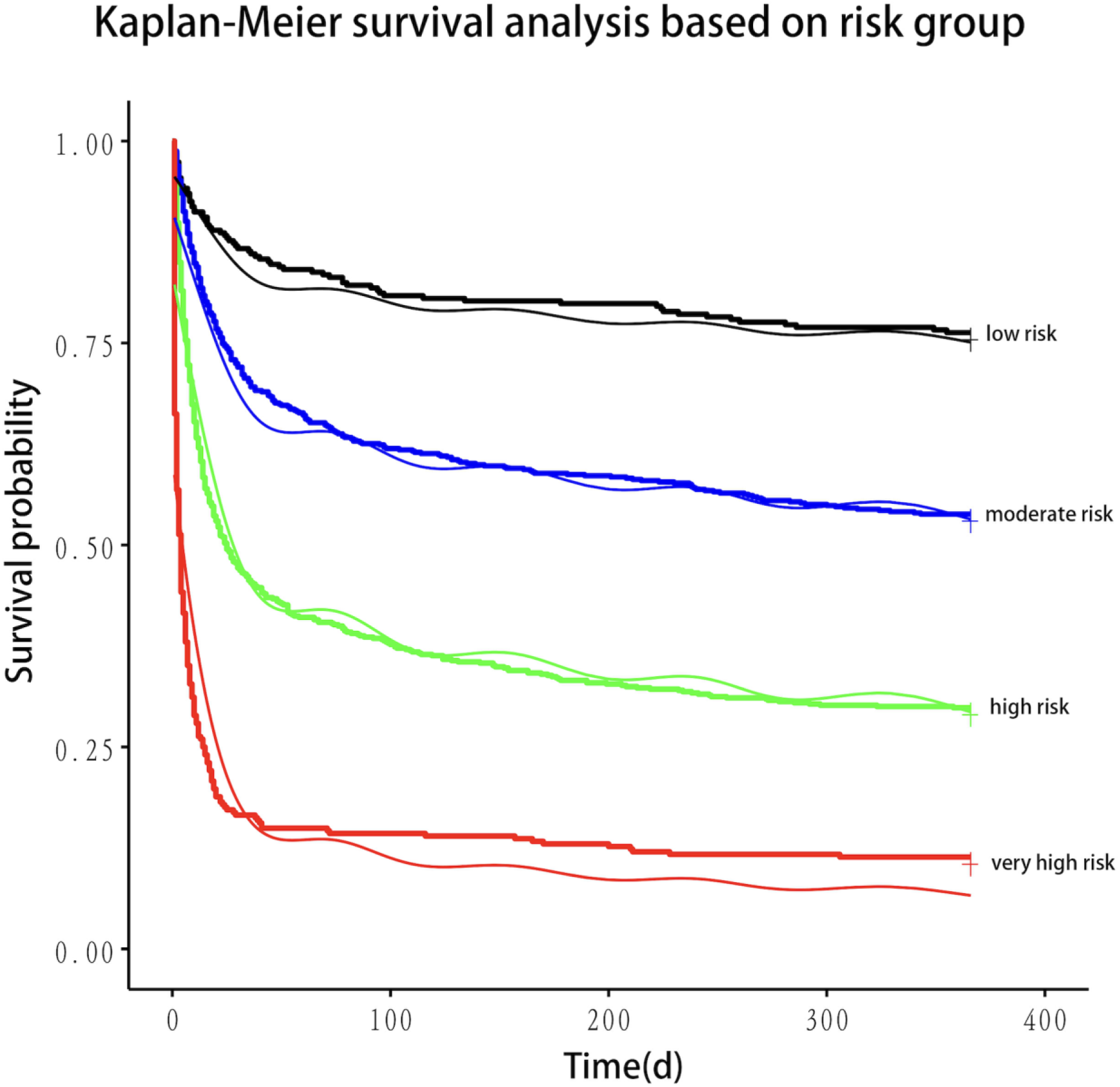
Kaplan-Meier plotter for risk stratification based on nomogram.

### Performance and Validation of the Model

The performance of the CSSN was evaluated using multiple metrics, including the c-statistic, AUC, calibration plots, and DCA. The c-statistic for the predictive model was 0.75 (95% CI 0.73–0.77) in the training cohort and 0.733 (95% CI 0.70–0.77) in the validation cohort, indicating good predictive accuracy. ROC curves for 30-day survival predictions yielded corresponding AUC values (Figure 4). In the training cohort, the AUC for CSSN predicting 30-day survival was 0.76, compared to 0.73 for the CSS. In the validation cohort, CSSN had an AUC of 0.73, while the CSS had an AUC of 0.72, demonstrating superior discriminative power by CSSN. Calibration plots (Figure 5, panels A and B) revealed strong agreement between nomogram-predicted and actual survival probabilities, indicating better model calibration than CSS. DCA also demonstrated the superior performance of the CSSN over the CSS (Figure 6, panels A and B).

**Figure 4.**
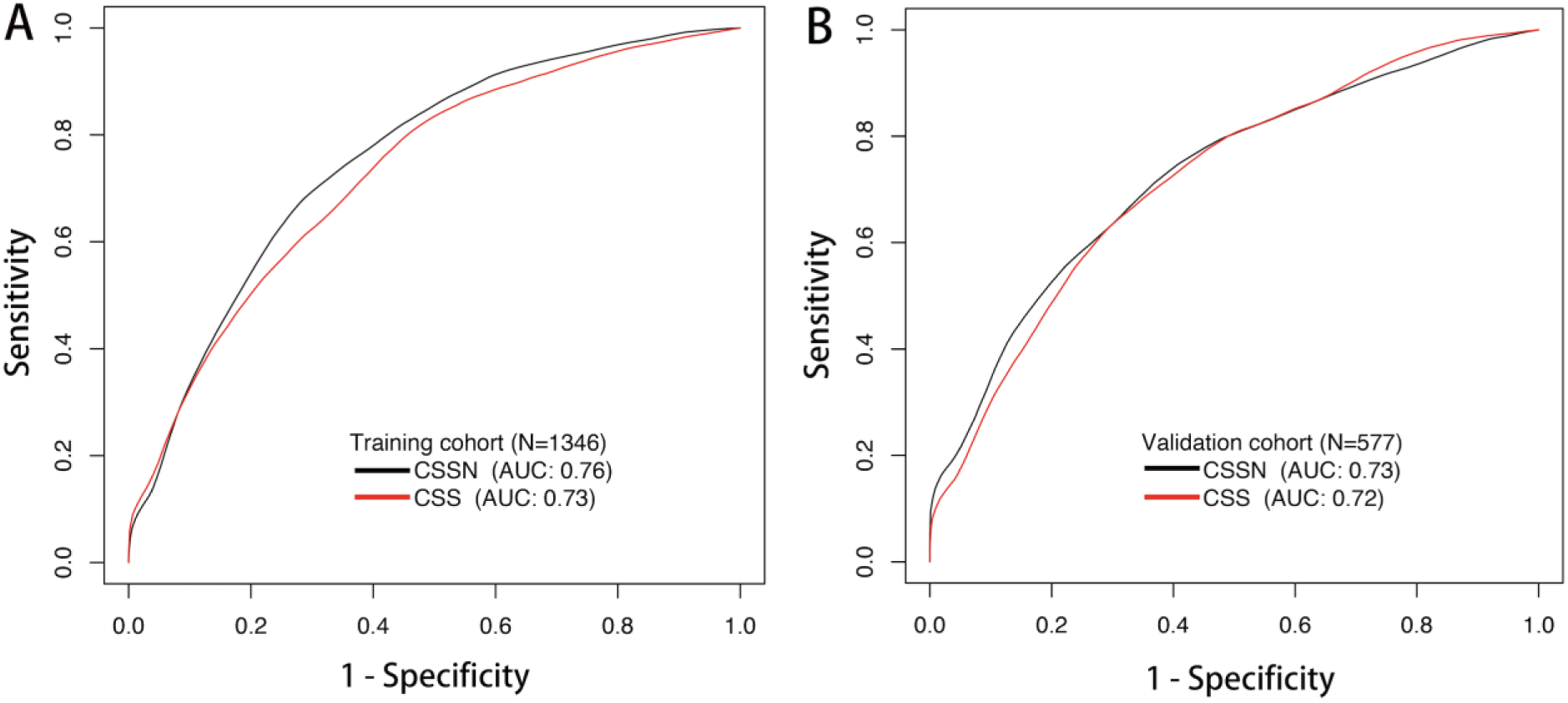
The time-dependent ROC curve and AUC of the CSSN and CSS in the training and validation cohort. Abbreviations: AUC, the area under the ROC curve; ROC, receiver operating characteristic. CSSN, Cardiogenic Shock Survival Nomogram; CSS, Cardiogenic Shock Score.

**Figure 5.**
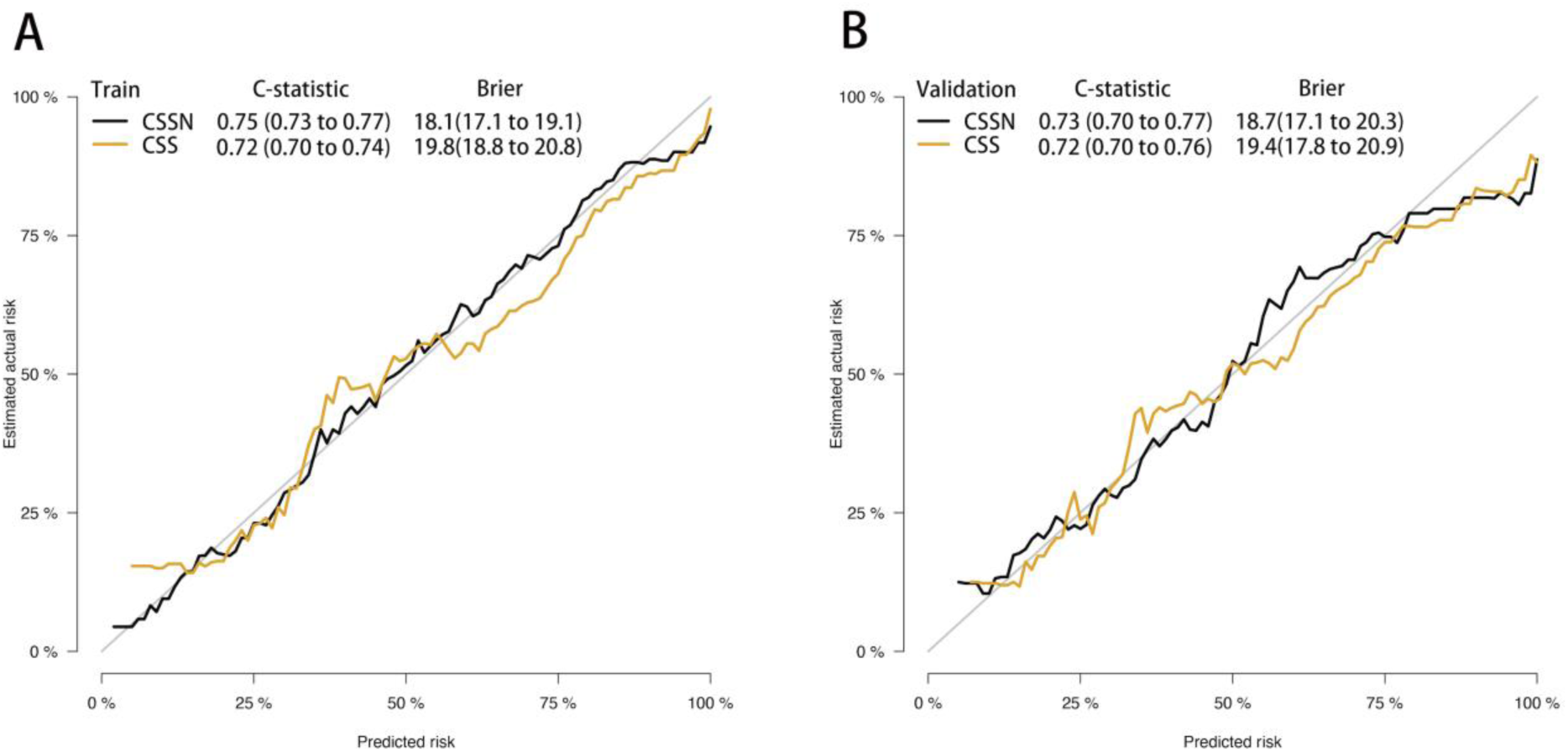
Calibration plot of the CSSN and CSS in the training and validation cohort. Panels A demonstrate the calibration plots for predicting 30-day survival in the training cohort, while panel B illustrates the calibration plots for predicting 30-day in the validation cohort.

**Figure 6.**
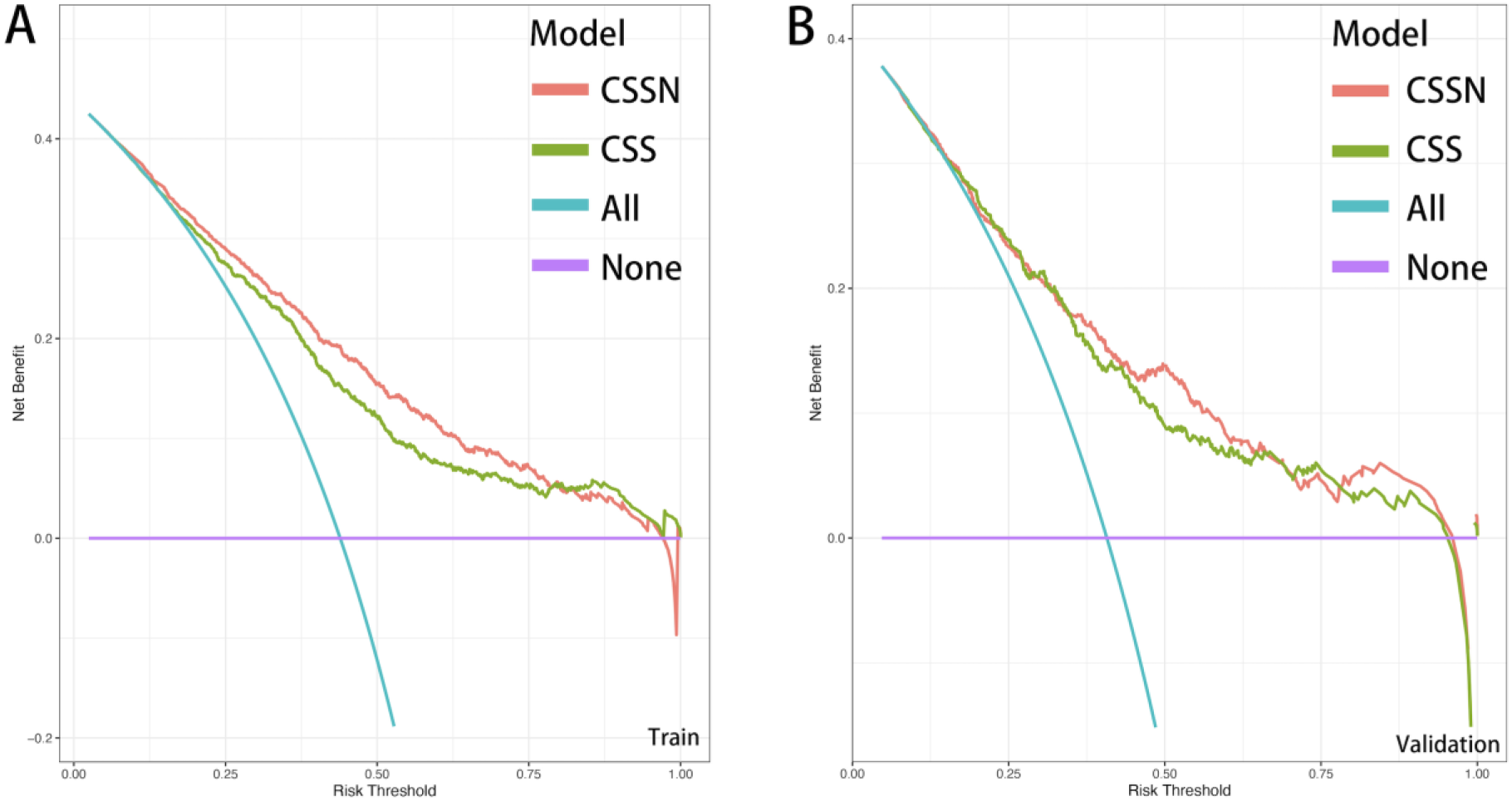
Decision curve analysis of CSSN and CSS in the training and validation cohorts.

### Sensitivity Analyses

Sensitivity analyses were conducted to verify the robustness of the model, as detailed in Table 3: (1) In the subset of patients with AMI-CS, the model demonstrated a c-statistic of 0.77 (95% CI: 0.74-0.80) and an AUC of 0.77 for CSSN. (2) In the subset of non-AMI-CS patients, the model showed a c-statistic of 0.73 (95% CI: 0.71-0.76) and an AUC of 0.73 for CSSN. These analyses confirmed the predictive capability and reliability of the nomogram across different patient subsets, with the highest accuracy observed in patients with AMI-CS.

**Table 3.** Sensitivity Analyses.

|  | **AMI cases** |  | **non-AMI cases** |  |
| --- | --- | --- | --- | --- |
| **Number** | **(N = 541)** |  | **(N = 1143)** |  |
| **c-statistic** | 0.768 (0.737, 0.798) |  | 0.734 (0.713, 0.755) |  |
| **AUC** | 0.768 |  | 0.732 |  |
| **Variables** | **HR (95%CI)** | **P** | **HR (95%CI)** | **P** |
| Age, year | 1.040 (1.029, 1.052) | <0.0001 | 1.030 (1.024, 1.037) | <0.0001 |
| SBP, mmHg | 0.980 (0.971, 0.989) | <0.0001 | 0.993 (0.988, 0.999) | 0.0161 |
| SpO<sub>2</sub>, % | 0.987 (0.976, 0.996) | 0.0167 | 0.983 (0.978, 0.989) | <0.0001 |
| Hemoglobin, g/dL | 0.929 (0.855, 1.009) | 0.0814 | 0.940 (0.893, 0.989) | 0.0164 |
| Serum creatinine, umol/L | 1.001 (1.001, 1.002) | 0.024 | 1.001 (1.001, 1.001) | 0.0008 |
| Blood glucose |  |  |  |  |
| <11.1 mmol/L | Reference |  | Reference |  |
| ≥11.1mmol/L | 1.530 (1.177, 1.989) | 0.0015 | 1.294 (1.067, 1.569) | 0.0088 |
| pH (Arterial blood gas) |  |  |  |  |
| <7.1 | Reference |  | Reference |  |
| 7.1~7.35 | 0.424 (0.258, 0.697) | 0.0007 | 0.535 (0.374, 0.766) | 0.0006 |
| >7.35 | 0.263 (0.141, 0.481) | <0.0001 | 0.525 (0.344, 0.780) | 0.0027 |
| Arterial lactate, mmol/L | 1.085 (1.051, 1.119) | <0.0001 | 1.078 (1.054, 1.103) | <0.0001 |
| Norepinephrine | 1.645 (1.178, 2.298) | 0.0035 | 2.012 (1.607, 2.519) | <0.0001 |

Multivariate Cox proportional hazards model for 30-day mortality in AMI and non-AMI cases.
Abbreviations: HR, hazard ratio; CI, confidence interval; SBP, systolic blood pressure; SpO<sub>2</sub>, peripheral oxygen saturation.

## Discussion

In this study, we developed a predictive model for assessing the survival rates of CS patients at 30-day intervals to evaluate the risk of death. Our model, constructed using easily accessible variables from the emergency setting, demonstrated better performance than CSS. Visualized as a Cardiogenic Shock Survival Nomogram (CSSN), our model is simple and user-friendly. Furthermore, this is the first prognostic model for cardiogenic shock mortality risk presented as a nomogram, enhancing its clinical applicability and facilitating timely and appropriate clinical decision-making.

### The Importance of Risk Assessment in CS Patients

Risk assessment in CS patients is crucial for guiding physicians in making more appropriate treatment strategies, particularly in deciding whether to use mechanical circulatory support (MCS). MCS can somewhat mitigate the issue of insufficient cardiac output in CS patients, but insufficient evidence remains to prove its efficacy in reducing mortality rates^2,17–21^. This is primarily due to the many complications associated with MCS use, such as vascular complications, thrombosis, limb ischemia, infection, and fatal hemorrhage^2,20,22^. Inappropriate use of MCS can lead to an imbalance of risks and benefits. For example: (1) Using MCS in low-risk patients may alleviate their insufficient cardiac output, but its complications might outweigh the benefits, potentially reducing the patient’s quality of life and survival rate. High-risk patients might be misclassified as low-risk, thereby missing the opportunity for timely MCS intervention, leading to adverse outcomes. (3) For patients with highly severe conditions (meager survival rates), using MCS may not provide enough survival benefits and could impose a heavy medical burden. Thus, timely and accurate risk assessment and appropriate treatment strategies are critical challenges clinicians face and are crucial to benefiting patients.

Risk assessment of CS patients also aids in guiding the implementation of clinical trials. Previously, several randomized trials investigating the use of MCS in CS patients failed to demonstrate survival benefits. For instance, the large randomized trial, IABP-SHOCK II, did not show that intra-aortic balloon pump (IABP) could lower the 30-day, 1-year, and 6-year mortality rates in patients with CS complicating acute myocardial infarction (AMI) undergoing early revascularization^17,21,23^. Similarly, previous ECMO and Impella trials failed to demonstrate survival improvements in CS^1,19,20,24^. However, the latest randomized trial published by Møller and colleagues in the New England Journal of Medicine (NEJM) has changed this perspective^25^. In this trial, 360 AMI-CS patients were randomly assigned to either the Impella or the standard treatment group, with all patients undergoing revascularization. Results showed that the 180-day mortality rate in the Impella group was 45.8%, significantly lower than the 58.5% in the standard treatment group (hazard ratio = 0.74; 95% CI 0.55-0.99; P = 0.04). This marks the first MCS device proven to reduce mortality in CS patients in an RCT since the 1999 SHOCK trial. Notably, this trial differed from previous ones: (1) The trial excluded patients in SCAI-CSWG stages A and B, avoiding the imbalance of risks and benefits from using Impella in low-risk patients. (2) Patients who had been resuscitated from out-of-hospital cardiac arrest and remained comatose upon arrival at the cardiac catheterization laboratory were excluded. This likely excluded extremely critical patients who might not benefit neurologically or survive from MCS. (3) The trial’s follow-up period was 180 days, whereas previous studies typically had 30-day follow-ups. The SHOCK trial indicated that PCI’s effect on 30-day mortality in AMI-CS patients was neutral, but benefits were seen at 180 days^26^. Therefore, 30 days as a primary endpoint may be too early to assess the intervention’s effects adequately. These findings highlight the importance of considering both short-term and long-term outcomes, not just short-term ones.

In summary, risk assessment and stratification of CS patients are essential for guiding physicians in patient management and facilitating the successful implementation of clinical trials. Developing a simple and easy-to-use predictive model that considers both short-term and long-term outcomes of CS will be of significant clinical value.

### Previous Models

Currently, three primary models are used to predict the prognosis of CS patients: IABP-SHOCK II risk score^7^, CardShock risk Score^8^, and Cardiogenic Shock Score (CSS)^9^. These models provide tools for the management of CS patients and the conduct of clinical trials. However, they have not been widely used in clinical practice, possibly due to limitations in population applicability, easy-to-use, and consideration of long-term outcomes. Population applicability may be the primary factor limiting the practical utility of these models. The IABP-SHOCK II risk score was developed for AMI-CS patients and requires post-PCI TIMI flow grades, making it unsuitable for non-AMI-CS patients. The CardShock score was developed in 2015 for all CS patients but included most AMI-CS patients (81%). In addition, the sample size was only 219, which may limit the applicability of the results to non-AMI-CS patients. Historically, AMI has been the primary cause of CS, constituting the majority of cases (> 80%)^8^. However, recent epidemiological surveys indicate that the proportion of AMI-CS in CS gradually decreases, now accounting for about 30%^3,12^. In the 2017-2018 North American CCCTN study, only 30% of CS cases were related to AMI, and among non-AMI-CS patients, approximately two-thirds had a history of heart failure^12^. Therefore, models unsuitable for non-AMI-CS patients may limit their clinical application.

Secondly, the simplicity of the model and its consideration of long-term outcomes also determine whether clinicians will use them in practice. The CSS, a recent score system developed to predict 30-day mortality in CS, has better population applicability than the IABP-SHOCK II and CardShock risk scores, covering all causes of CS^9^. The CSS includes nine easily obtainable parameters (age, sex, AMI-CS, systolic blood pressure, heart rate, pH, lactate, glucose, and cardiac arrest) and demonstrates good accuracy (c-statistic = 0.74). However, the CSS only focuses on the 30-day mortality of CS patients and does not consider long-term outcomes. In reality, the recovery of CS patients within one year after discharge is not optimistic, and survival rates continue to decline^4,27^. Currently, the in-hospital mortality rate of CS patients is about 30-40%, and the one-year mortality rate can reach 50-60%^3^. Once again, the latest Impella trial successfully demonstrated that the Impella microaxial pump could reduce mortality in CS patients, and the follow-up period for this trial was 180 days (6 months) rather than 30 days^25^. This indicates that the long-term outcomes of CS patients are also worthy of our attention. This study compared the performance of the CSS and the newly developed CSSN using c-statistic, time-dependent ROC, calibration plots, and decision curve analysis (DCA), indicating higher accuracy of the CSSN. Furthermore, the study employed CSSN to predict 30-day mortality risk for all patients, grouping them based on risk levels. Significant differences were observed in survival curves among the groups over one year.

### CSSN and Its Clinical Contributions

A growing body of evidence suggests that several clinical indicators can be used to predict the severity and outcomes of patients with CS^7–9,28^. Based on the previous model, the CSSN was developed, including the IABP-SHOCK II risk^7^, CardShock risk score^8^, and CSS^9^. In this study, we used LASSO regression and the multivariable Cox model to screen and confirm easily obtainable parameters in emergency settings and those widely recognized in previous studies. Consequently, age, SBP, SpO2, hemoglobin, serum creatinine, blood glucose, pH, arterial lactate, and norepinephrine were identified and used to develop the prognostic nomogram in our study. This nomogram demonstrated good discrimination and calibration in predicting patients’ 30-day probability of survival with CS, as assessed by the c-statistic, AUC value, calibration plots, and clinical decision curve analysis indicating good performance and high value for clinical use.

Based on previous models and after multiple refinements and optimizations, our nomogram has several advantages^7–9^: (1) Comprehensive Application: Our model is designed for CS resulting from any cause, and we have confirmed its accuracy in both AMI-CS and non-AMI-CS through sensitivity analysis. (2) Simplicity and Accessibility: Our model includes only nine parameters that are easily accessible in emergency settings, and it is presented in the form of a nomogram, making it simple and easy to use. Clinicians can quickly assess a patient’s risk using our nomogram. (3) Holistic Approach: Our model performs well even one year after discharge, enabling clinicians to provide more appropriate treatment measures by weighing risks and benefits.

Furthermore, our model can be used with SCAI classification, allowing physicians to assess a patient’s survival rates accurately. SCAI divides cardiogenic shock into five stages (A-E), corresponding to “at risk” for CS, “beginning” shock, “classic” CS, “deteriorating”, and “extremis”, respectively^12^. This staging is based on a comprehensive evaluation of blood pressure, heart rate, lactate levels, urine output, serum creatinine, vasopressor dose and duration, and blood pressure response to vasopressors. Through SCAI classification, clinicians can roughly understand the patient’s disease progression stage and provide appropriate treatment strategies^6^. Our nomogram can complement SCAI classification for a more precise assessment. For example, age is a critical factor influencing the prognosis of CS patients, and patients at the same SCAI stage may face different risks due to age differences^29,30^. Combining CSSN with SCAI staging can enable a more precise assessment. Moreover, our study incorporates continuous variables into the model as much as possible to achieve more refined predictive capabilities, greatly enhancing its usability.

### Limitations

Despite the large sample size and thorough evaluation and validation, our study has several limitations inherent to retrospective research. The primary limitation is the inability to rule out unknown confounding factors, a common issue in retrospective studies. These unknown factors could potentially influence our predictive model’s outcomes and accuracy. Secondly, the CSSN is based on the MIMIC database, and all patients received ICU hospitalization, which may introduce bias when applied to patients in other regions. Additionally, because the MIMIC database does not include etiological diagnoses of CS, we could not obtain specific causes of CS in patients; the diagnosis of AMI-CS was based on the admission diagnosis. Despite these limitations, we believe the CSSN remains a valuable tool for clinicians in managing cardiogenic shock. The model’s ability to predict outcomes of CS, combined with its simplicity, makes it a practical addition to current clinical decision-making processes. Future prospective studies and validation in diverse patient populations are needed further to enhance the robustness and applicability of the CSSN.

## Conclusion

In this study, we developed a prognostic model for predicting survival rates in patients with cardiogenic shock, known as the Cardiogenic Shock Survival Nomogram (CSSN). The CSSN is constructed using easily obtainable parameters in an emergency setting. This model is straightforward, intuitive, and easy to use. Utilizing the CSSN aids physicians in the early risk assessment of CS patients and helps formulate targeted treatment strategies, potentially improving patient outcomes.

## Data Availability

All data generated or analyzed during this study have been included in this published article. The raw data are available in the physionet (https://physionet.org/content/mimiciv) after completing the appropriate training and registration

## Acknowledgments

The authors thank the original study group for providing data for the current analysis.

## Consent for publication

Not applicable.

## Ethics approval and consent to participate

The collection of patient information and creation of the research resource underwent a review by the Institutional Review Board at the Beth Israel Deaconess Medical Center. A waiver of informed consent was granted, and the data-sharing initiative was approved. As a result, the ethical approval statement and the need for informed consent were waived for this article. All methods conducted in this study adhered to relevant guidelines and regulations, including the principles outlined in the Declaration of Helsinki.

## Availability of data and materials

All data generated or analyzed during this study have been included in this published article. The raw data are available in the physionet (https://physionet.org/content/mimiciv) after completing the appropriate training and registration^14–16^.

## Nonstandard Abbreviations and Acronyms

CS: Cardiogenic Shock
CSS: Cardiogenic Shock Score
CSSN: Cardiogenic Shock Survival Nomogram
LASSO: Least Absolute Shrinkage and Selection Operator
MCS: mechanical circulatory support
MIMIC-IV: Medical Information Mart for Intensive Care IV
SCAI: Society for Cardiovascular Angiography and Intervention

